# Quarantine and testing strategies to reduce transmission risk from imported SARS-CoV-2 infections: a global modelling study

**DOI:** 10.1101/2021.06.11.21258735

**Authors:** Billy J Quilty, Timothy W Russell, Samuel Clifford, Stefan Flasche, Suzanne Pickering, Stuart JD Neil, Rui Pedro Galão, W John Edmunds, CMMID COVID-19 Working Group

**Affiliations:** Centre for Mathematical Modelling of Infectious Diseases, London School of Hygiene & Tropical Medicine, LONDON WC1E 7HT, United Kingdom; Department of Infectious Diseases, School of Immunology & Microbial Sciences, King’s College London, LONDON SE1 9RT, United Kingdom

## Abstract

**Background:** Many countries require incoming air travellers to quarantine on arrival and/or undergo testing to limit importation of SARS-CoV-2.

**Methods:** We developed mathematical models of SARS-CoV-2 viral load trajectories over the course of infection to assess the effectiveness of quarantine and testing strategies. We consider the utility of pre and post-flight Polymerase Chain Reaction (PCR) and lateral flow testing (LFT) to reduce transmission risk from infected arrivals and to reduce the duration of, or replace, quarantine. We also estimate the effect of each strategy relative to domestic incidence, and limits of achievable risk reduction, for 99 countries where flight data and case numbers are estimated.

**Results:** We find that LFTs immediately pre-flight are more effective than PCR tests 3 days before departure in decreasing the number of departing infectious travellers. Pre-flight LFTs and post-flight quarantines, with tests to release, may prevent the majority of transmission from infectious arrivals while reducing the required duration of quarantine; a pre-flight LFT followed by 5 days in quarantine with a test to release would reduce the expected number of secondary cases generated by an infected traveller compared to symptomatic self-isolation alone, R_s_, by 85% (95% UI: 74%, 96%) for PCR and 85% (95% UI: 70%, 96%) for LFT, even assuming imperfect adherence to quarantine (28% of individuals) and self-isolation following a positive test (86%). Under the same adherence assumptions, 5 days of daily LFT testing would reduce R_s_ by 91% (95% UI: 75%, 98%).

**Conclusions:** Strategies aimed at reducing the risk of imported cases should be considered with respect to: domestic incidence, transmission, and susceptibility; measures in place to support quarantining travellers; and incidence of new variants of concern in travellers’ origin countries. Daily testing with LFTs for 5 days is comparable to 5 days of quarantine with a test on exit or 14 days with no test.

## Introduction

SARS-CoV-2 emerged in late 2019 in Wuhan, China, and spread rapidly around the world through international travel. Many countries have since acted to reduce importation of infectious individuals with measures including border closures, partial travel restrictions, entry or exit screening, and quarantine of travellers (1). The effectiveness of such policies in detecting infectious travellers will be influenced by factors including the natural history of SARS-CoV-2 infection, the performance of tests used for screening, and the level of adherence with guidelines for both quarantine (i.e. the separation of individuals, independent of their case-status, from the wider community) and self-isolation (i.e. the separation of symptomatic or test-positive individuals from the wider community) (2). In addition, the absolute risk of infected entries to each country will be determined by the volume of travel from, and prevalence of infection in, other countries(3). The emergence of more transmissible and virulent variants of SARS-CoV-2 in late 2020 has brought a new impetus to assessing the effectiveness of travel restrictions. Here we combine previously published models for estimating prevalence and incidence, travel volume, infectivity, and effectiveness of testing and quarantine strategies to estimate the reduction in risk of importing new infections and compare the number of infectious arrivals to the size of the domestic epidemic in 99 countries. We also estimate the extent to which each strategy (quarantine of varying duration with/without test at exit; daily LFTs; and the use of pre-flight tests) may avert onward transmission in the destination country by considering the reduction in the expected reproduction number of arrivals compared to a scenario with no interventions specifically targeting travellers.

## Method

Briefly, we estimate the proportion of infected travellers who would be detected by each of the considered quarantine and testing scenarios (and at which point in their journey, Figure 1) and calculate the relative reduction in the expected number or secondary cases generated by an infected traveller, *R*. We then estimate the total number of travellers arriving to each destination country and the prevalence of SARS-CoV-2 infection in each origin country in order to estimate the number of infected travellers who would arrive under a no-intervention scenario. We then describe the risk of importation as being the ratio of daily infectious arrivals and domestic incidence in the destination country, and explore exportation risk for the variant of concern B.1.1.7 from the UK as a case study. The following subsections describe each component of the model in further detail.

**Figure 1:**
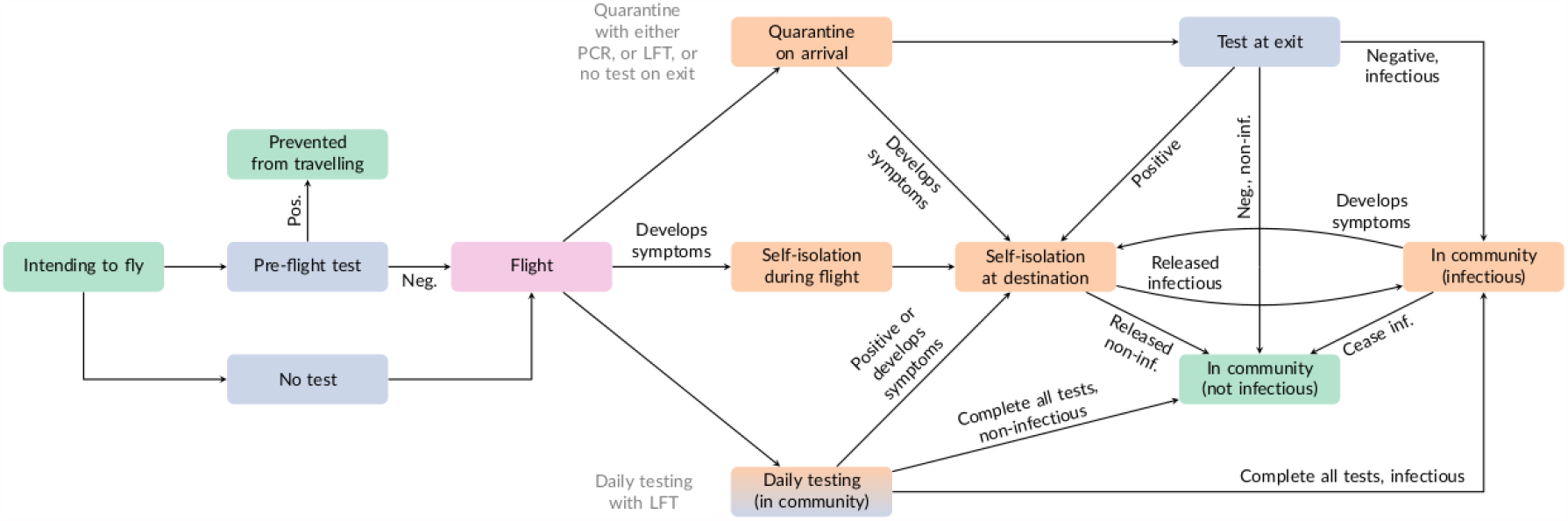
Schematic of quarantine and testing strategies, showing endpoints of travel process (green), flight (red), pre- and post-flight testing (blue), and quarantine and self-isolation points (orange). Individuals are assumed to only develop symptoms once. Individuals are subject to a strategy of either post-flight quarantine (upper arm) or daily testing (lower arm); under both types of strategy, a positive test or development of symptoms results in self-isolation and eventual release either while still infectious or while not infectious. Adapted from (4).

### Viral load trajectories

For each infected individual, we simulate a viral load trajectory based on publicly available data of the longitudinal sampling of individuals by Polymerase Chain Reaction (PCR) (5,6) from which we derive individual-level infectiousness over the course of infection and the probability of testing positive at time of sampling (Figure 2). Key assumptions of viral load dynamics are that both symptomatic and asymptomatic infections’ cycle threshold (Ct) values peak, in accordance with the incubation period, 5.1 days (95% range: 2.3, 11.5 days) after the infecting exposure (7), at a mean Ct of 22.3 (SD: 4.2) (5). We assume that viral shedding continues until 17 days (SD: 0.94 days) after exposure for symptomatics (6) and that for asymptomatics this duration is reduced by 40% (5,6). We also assume that between 24% and 38% of individuals are asymptomatic throughout the course of infection (8), and, for each model run, sample the fraction of intending travellers who are infected from a Beta distribution whose 95% interval matches (24%, 38%). Assuming that the ability to culture virus is a reasonable proxy for infectiousness (9) we fitted a logistic regression model to estimate the probability of infectiousness from the ability to culture virus for a given viral load (in Ct) using data from Pickering et al. (10) using R’s *glm* function (Figure 2B). This model is then used to estimate an instantaneous probability of infectiousness at each timepoint in the course of infection given the previously simulated Ct values (Figure 2C). Integrating under the entirety of this infectiousness curve gives an individual-level value of infectiousness which is proportional to *R*_0_ (11). This is then censored on the left by the time of departure from the origin country to allow for the possibility of in-flight transmission. The infectiousness distribution can then be further truncated by quarantine or self-isolation upon symptom onset or a positive test result. The duration of the infectious period is defined as the period of time when Ct values are below 25, i.e, the approximate viral load at which the virus becomes culturable (Figure 2). Reduction in infectiousness is characterised as proportional, so the exact value of *R*_0_ need not be estimated or assumed.

**Figure 2:**
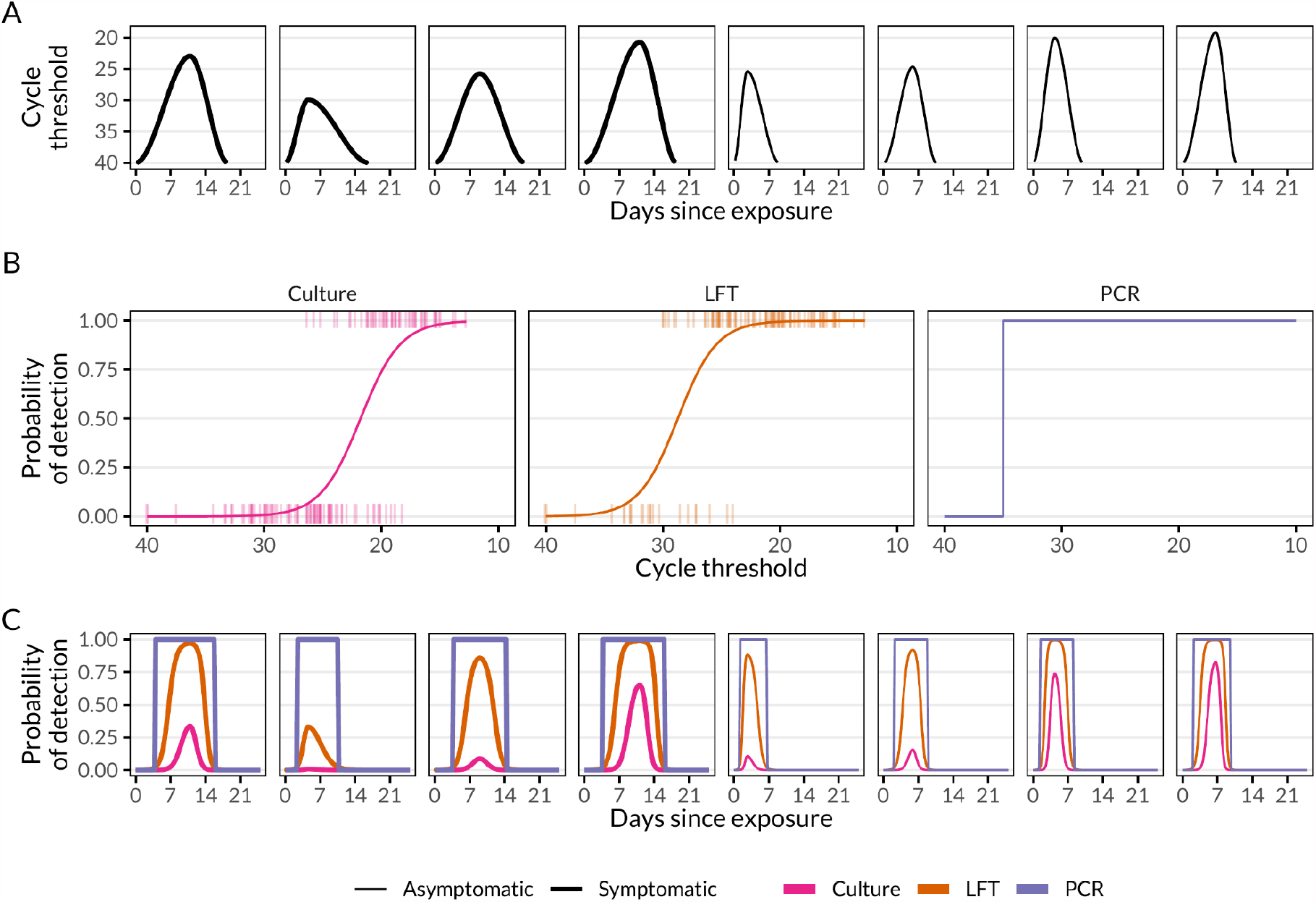
A) Eight randomly sampled viral load trajectories over the course of infection (symptomatic and asymptomatic), B) the functions to estimate probability of detection by culture (the assumed infectivity function), the Innova lateral-flow antigen test, and PCR, given a certain cycle threshold, and C) the produced detection curves over the course of infection.

### Testing

We model the sensitivity of LFTs using the results of the Innova SARS-CoV-2 Antigen Rapid Qualitative Test as evaluated by Pickering et al. (10) namely whether a swab returned a positive result under LFT when infection was detected by PCR with an associated Ct value (Figure 2). We fitted a logistic regression model using *glm* in R to estimate the mean probability of detection for a given Ct value, i.e, the viral load at time of sampling, after regrouping “strongly positive”, “clearly positive”, and “weakly positive” as “positive” test results (Figure 2B). PCR tests are assumed to have a detection threshold at a Ct value of 35; below this threshold value, we assume PCR is 100% sensitive and that it is 0% sensitive for Ct values greater than 35 (Figure 2B). Sensitivity of LFTs varies with viral load, and therefore with time. At a Ct of 35 the probability of detection is 0.02; at the mean simulated peak Ct of 22.3 the probability of detection is 0.98. The proportion adhering to self-isolation following a positive test, either by PCR or LFT, is assumed to be 86% (12).

### Intervention strategies

Air travellers may be subjected to interventions that vary in stringency with the aim of both preventing infectious arrivals and reducing their potential for transmission in the destination country (Figure 1). Within our modelling framework, travellers may either not be tested (baseline) or be tested pre-flight by either PCR or LFT, where a negative test result is required to board the plane (with sensitivity analysis for varying delays from test to departure). Upon arrival (an assumed six hours after departure) travellers are subject to quarantine and/or testing (Figure 1). Travellers may enter a post-flight quarantine of 0 (baseline), 3, 5, 7, 10 or 14 days duration with, on the final day of quarantine, either no test, an LFT test, or a PCR test. A positive test at the end of quarantine leads to an additional 10 days of self-isolation. Based on published estimates from Norway, we assume that the proportion of individuals adhering to quarantine in the absence of symptoms is 28%, with individuals either fully adherent or non-adherent, although asymptotically there should be no difference between 28% of individuals adhering and all individuals adhering 28% (13). Alternatively, travellers may take LFT tests daily for 3, 5, 7 or 10 days without quarantine, only self-isolating upon the receipt of a positive test. We assume that individuals who develop symptoms (fever or high temperature, a cough that has lasted for at least several hours, shortness of breath, aches and pains (e.g. in back, neck, shoulders or joints), blocked nose, sore throat and feeling unusually tired) will self-isolate for 10 days (or not board the plane if symptomatic upon flight departure), with the proportion adhering to symptomatic self-isolation being 71% as reported by the ONS in the UK (14).

### Detection process

Infected travellers may be detected either: prior to departure by either a pre-flight PCR test three days prior to departure or an LFT test immediately pre-flight; by a post-flight test (either daily LFT testing or at the end of quarantine); or by developing symptoms either during their flight or during or after the quarantine period (each of which triggers the need for isolation). All passengers detected pre-flight are considered to contribute no risk of further infection in the destination. Based on the viral load dynamics of those who do travel, we estimate whether individuals still pose a risk of community transmission in the destination country. Those who still pose a risk are either: asymptomatic, infectious but not detected by daily testing; those who do not adhere to quarantine guidance while infectious; those who remain infectious after their release from quarantine (either without a test or a false negative result); those who do not isolate on development of symptoms; or those who do not isolate after receiving a positive test result. For each quarantine and testing scenario we simulate 100 infected travellers (bootstrapped 100 times to estimate uncertainty) and estimate the proportion of travellers detected at each stage, whether they pose a risk of community transmission, and the expected reduction in *R*_0_ of each strategy. We also estimate the effectiveness of the best case for each strategy, i.e. the proportion adhering to quarantine, self-isolation after symptom onset, and self-isolation after a positive test, are all 100%, rather than 28%, 71% and 86% respectively. Results are presented, both for proportion of the would-be infectious or eventually infectious arrivals in each detection and infection group and the effectiveness of an intervention as a reduction in R, as medians and uncertainty intervals (UIs) calculated as the quantiles of the sampled values within the simulations for each scenario.

### Estimating the number of travellers

We estimate the number of infectious arrivals to a given country by the method of Russell et al. (3). By applying an estimate of case under-ascertainment rates based on reported deaths and infection fatality ratio, we estimate the prevalence of infection in each country. We assume that the prevalence in the general population is the same as the prevalence in travellers and so the total number of infectious arrivals per day for a given country is the sum of the estimated daily travel volume from each origin to that country, weighted by estimated prevalence. Estimates of travel volume are obtained from the publicly available OpenSky database, which provides daily data on the number of flights between airports (15). We estimate the number of arrivals per flight (at the current stage of the pandemic) with the mean number of travellers per flight, 142, the total number of passenger movements divided by the total number of passenger flight movements as of 20 March 2021 (16) and note that this implies similar aircraft are used on all international flights. Countries with no available flight data (n=71) were excluded from the model. The OpenSky database is estimated to cover 45% of the total number of flights globally, with methods and explicit coverage estimates detailed in Strohmeier et al. (17).

### Variants of concern

As a case study to investigate the importation risk of SARS-CoV-2 Variants Of Concern (VOCs) relative to domestic incidence, we estimated the number of infectious cases exported from the United Kingdom infected with the B.1.1.7 Variant of Concern (35) to six other countries. Total domestic incidence (calculated using the underascertainment model described previously) in destination countries was used as the denominator as local B.1.1.7 incidence data were not available. The choice of B.1.1.7 was made due to data availability (both for the variant and flights) but the approach is variant agnostic. We show results for two countries selected at random from each of High Income Countries and Upper Middle Income Countries (according to World Bank classifications) as well as the United States of America and Singapore as countries with, respectively, high and low incidence.

All analyses were conducted in R version 4.0.5 (18).

## Results

### Outcome of quarantine and testing for infectious or eventually infectious arrivals

Pre-flight LFT and PCR testing detects 66% (95% UI: 48%, 86%) and 85% (95% UI: 73%, 96%) of infectious and eventually infectious travellers, respectively, indicating that pre-flight testing alone may play a substantial role in preventing the seeding of new outbreaks by preventing their arrival in the destination country (Figure 3).

**Figure 3:**
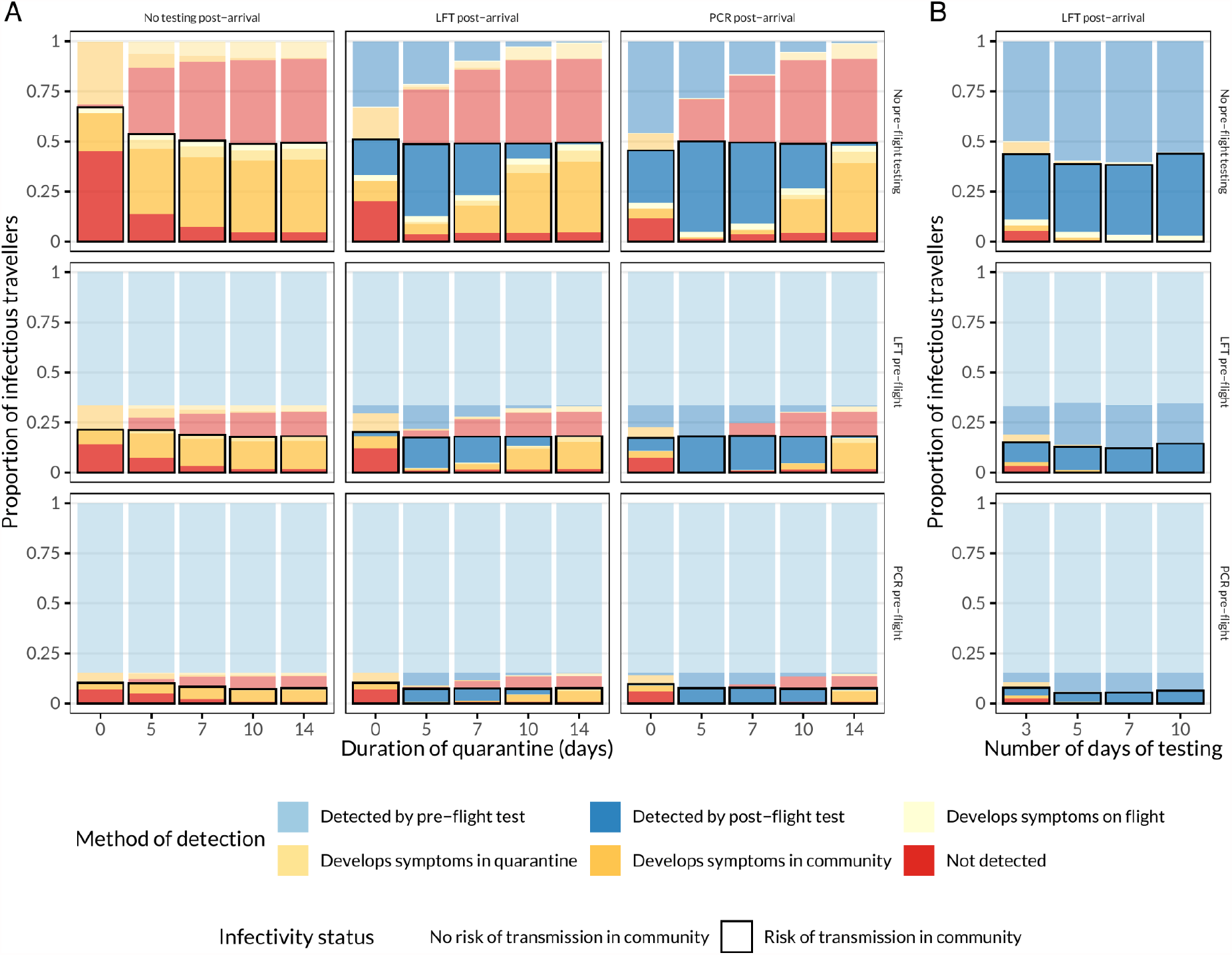
Mean proportion of intending travellers who would otherwise arrive infectious or become infectious detected at each stage of the quarantine and testing strategies (either: A) quarantine with no test, Lateral Flow test, or Polymerase Chain Reaction test or B) Daily Lateral Flow testing, and) assuming 28% adherence to quarantine guidance, 71% adherence to isolation guidance after the onset of symptoms and 86% adherence to isolation guidance after receipt of a positive test. Solid colours with black borders are those individuals who pose a risk of transmission in the community (i.e, still in the infectious period after leaving quarantine, self-isolation, or finishing the testing programme); semi-transparent colours pose no risk of transmission in the community.

In the baseline scenario of imperfect adherence (28% adherence to quarantine guidance, 71% adherence to isolation guidance after the onset of symptoms and 86% adherence to isolation guidance after receipt of a positive test), the most lenient travel restrictions (no quarantine and no pre-flight test) results in 67% (95% UI: 40%, 92%) of infectious or eventually infectious travellers entering the country undetected (Figure 3). This group is comprised of 7% (95% UI: 5%, 11%) of arrivals who develop symptoms in flight and do not self-isolate, 20% (95% UI: 7%, 38%) who develop symptoms on arrival and do not self-isolate, and 45% (95% UI: 20%, 66%) asymptomatic infections. With increasing duration of quarantine, those who become symptomatic will develop symptoms while in quarantine instead of in the community, and those who are released from quarantine are more likely to no longer be infectious. The addition of PCR or LFT testing at the conclusion of quarantine periods results in additional detection, allowing for their isolation and release when likely no longer infectious (Figure 3). After 5 days of quarantine with no pre-flight test, PCR testing at the end of quarantine results in 49% (95% UI: 31%, 70%) of infectious or eventually infectious individuals entering the community, using an an LFT test instead results in a similar proportion (51% (95% UI: 32%, 71%)). The marginal benefit of a post-quarantine test-to-release is reduced for longer quarantines, as individuals are more likely to develop symptoms before the test is conducted, or become less likely to test positive as viral shedding wanes.

Under daily testing scenarios, travellers are released into the community upon arrival and are tested every day with lateral flow tests. After 5 days of testing, the proportion of cases which go undetected by testing or by the development of symptoms is 7% (95% UI: 5%, 13%), decreasing to 0% after 10 days of testing. The reduction in the number of people self-isolating due to the onset of symptoms also decreases with a greater number of tests as a result of individuals receiving positive tests before symptom onset. However, imperfect adherence to self-isolation following a positive test or the onset of symptoms substantially decreases the programme’s effectiveness, with 39% (95% UI: 18%, 55%) of infectious or eventually infectious arrivals posing a transmission risk in the community (Figure 3).

Sensitivity analysis (Figure S1) indicates that full adherence to quarantine and isolation guidelines substantially reduces the risk of individuals entering the community while infectious or incubating. The proportion of infectious arrivals decreases to 0% for a 14 day quarantine without a test and can be shortened with the same prevention of infectious arrivals achieved by the addition of a PCR test to release on Day 7 or an LFT on Day 10. A combination of pre-flight lateral flow or PCR tests (which may prevent 66% (95% UI: 48%, 86%) and 85% (95% UI: 73%, 96%) of infectious arrivals respectively) with a 7 or 5 day quarantine with full adherence with a PCR or lateral flow test to release, respectively, would also eliminate the risk of infectious arrivals (Figure S1).

Stratifying by whether the individual remains asymptomatic or first engages with the quarantine and testing symptoms while in a pre-symptomatic state indicates that longer quarantines ensure that asymptomatic infections exit quarantine no longer infectious (Figure S2). The effect of post-quarantine testing is to divert a substantial number of asymptomatic infections to self-isolation, where imperfect quarantine adherence results in increased transmission risk (Figure 6). The effect of pre-flight tests is to more than halve the number of infectious arrivals who may cause onwards transmission. As post-flight testing becomes more sensitive to low viral loads (early in infection and asymptomatic cases) by moving from no test to LFT to PCR, a greater number of infectious travellers are diverted to self-isolation, peaking around day 5 for both LFT and PCR in both asymptomatic and ever-symptomatic infections. Earlier post-quarantine tests are likely to miss early stage infections and later tests allow symptoms to develop in ever-symptomatic cases. For daily testing, pre-flight testing picks up as many cases as for quarantine (as this is a measure in the country of departure). In the absence of pre-flight testing, 3 days of daily testing still allows some infectious asymptomatic cases to go undetected (15% (9%, 35%)), the risk of which drops to zero by 10 days of testing. As viral load increases prior to onset of symptoms, detectability of a pre-symptomatic infection with a daily LFT means the development of symptoms is rare and the individual may be diverted to self-isolation with an adherence level of 86%.

### Transmission potential of arrivals

We estimate that individuals isolating only upon the onset of symptoms after they arrive (assuming an imperfect adherence of 71%) would on its own reduce *R*_0_ of arrivals by 45% (95% UI: 28%, 64%) (Figure S3). After adjusting for the assumption that symptomatic self-isolation is adhered to at this proportion in all scenarios (thus reducing *R*_*0*_ to *R*_*s*_), a pre-flight lateral flow test would reduce *R*_*s*_ by an additional 62% (95% UI: 39%, 88%) (Figure 4) and a pre-flight PCR test would reduce *R*_*s*_ by an additional 80% (95% UI: 64%, 96%). Testing immediately after the flight with no quarantine (assuming no pre-flight testing) results in a smaller reduction in *R*_*s*_ to pre-flight testing (LFT: 33% (95% UI: 15%, 53%), PCR: 51% (95% UI: 35%, 77%) (Figure 4), due to assumed lower adherence to post-positive test self-isolation in the destination country compared to the complete elimination of transmission of a positive pre-flight test.

**Figure 4:**
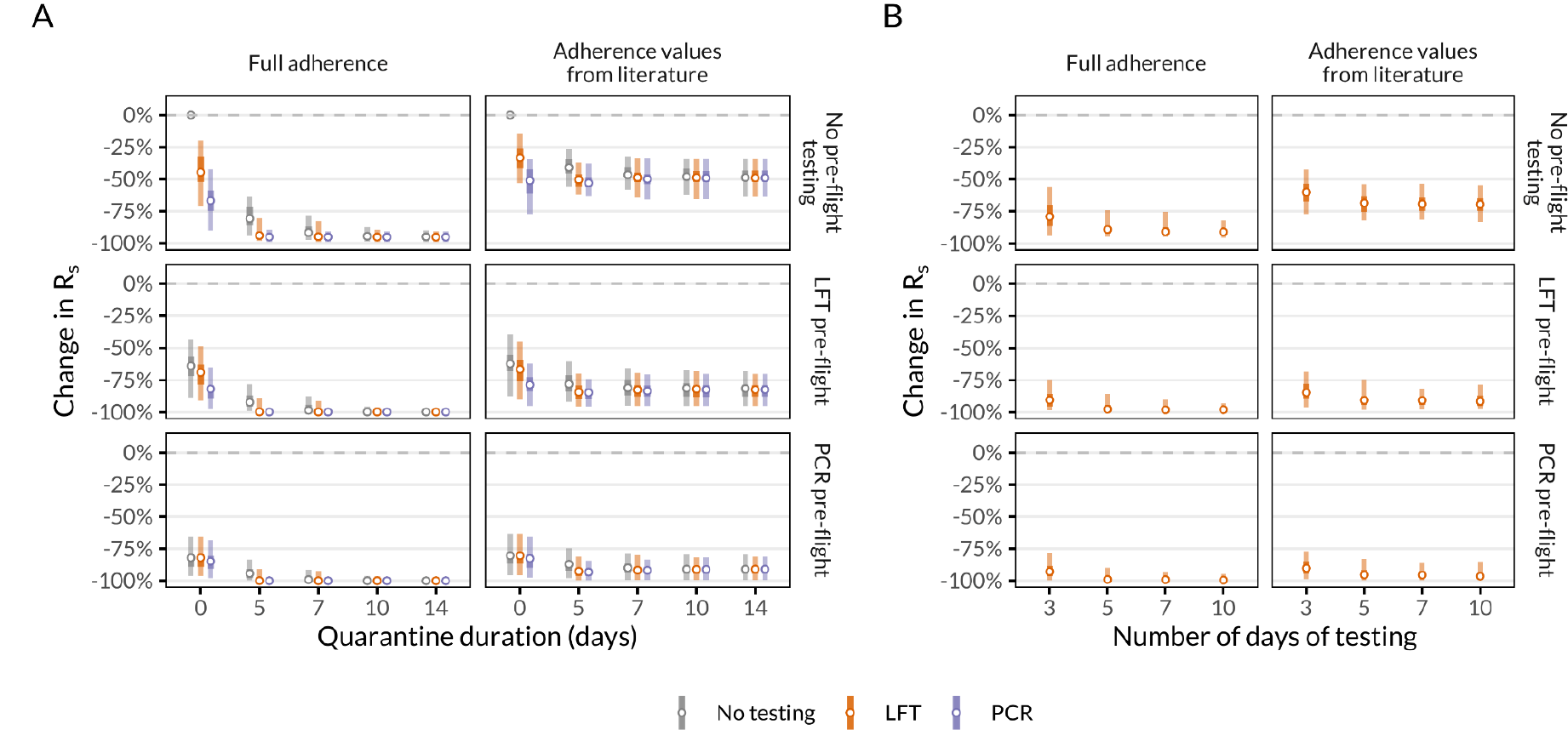
Change in R_s_ of infectious arrivals entering the community compared to symptomatic self-isolation only with full adherence (top row of plots) or adherence values from literature (28% of individuals adhering to quarantine and 86% adhering to post-positive test isolation, bottom row of plots), and with or without pre-flight tests. A) Quarantine of varying durations with or without testing with LFTs and PCR. B) Daily testing without quarantine with lateral flow tests, with self-isolation only upon a positive test result. Vertical lines represent 95% (outer) and 50% (inner) uncertainty intervals around medians (points). Note discrete x-axis values for quarantine duration and number of days of testing. Change in *R*_0_ (i.e, without adjustment for symptomatic self-isolation) shown in Figure S3.

Requiring travellers to quarantine upon arrival averts additional transmission, as does requiring a test at the end of quarantine. A 14 day quarantine would reduce *R*_*s*_ by 49% (95% UI: 34%, 64%) compared to symptomatic self-isolation alone, with identical impact with or without a test (Figure 4). If more individuals are assumed to adhere to self-isolation following a positive test than they would to quarantine (in the absence of symptoms), shorter quarantines of 5 days with a test to release would reduce *R*_s_ to a similar or greater degree (LFT: 50% (95% UI: 37%, 62%), PCR: 53% (95% UI: 38%, 63%) than that of a 14 day quarantine (Figure 4).

Combining both a pre-flight test and a short quarantine (e.g 5 days) on arrival with a test to release would reduce *R*_*s*_ further (Figure 4). A pre-flight LFT and 5 days of quarantine with an LFT to release would reduce *R*_*s*_ in would-be arrivals by 85% (95% UI: 70%, 96%); a pre-flight LFT with 5 days of quarantine with PCR to release would reduce *R*_*s*_ by 85% (95% UI: 74%, 96%); a pre-flight PCR with 5 days of quarantine with LFT to release would reduce *R*_*s*_ by 93% (95% UI: 81%, 100%); and a a pre-flight PCR with 5 days of quarantine with PCR to release would reduce *R*_*s*_ by 93% (95% UI: 84%, 100%).

Alternatively, replacing the requirement to quarantine with daily rapid tests upon arrival may reduce *R*_*s*_ by 60% (95% UI: 43%, 77%) for 3 days of testing, 68% (95% UI: 54%, 82%) for 5 days of testing, 69% (95% UI: 53%, 81%) for 7 days of testing, and 69% (95% UI: 54%, 83%) for 10 days of testing, assuming adherence to self-isolation is 86%. Combining daily testing with pre-flight testing may further reduce *R*_*s*_ (pre-flight LFT plus 5 days of tests on arrival: 91% reduction (95% UI: 75%, 98%); pre-flight PCR plus 5 days of tests on arrival: 95% reduction (95% UI: 83%, 100%) (Figure 4)).

If, instead of the published values, adherence is assumed to be 100% for both quarantine and self-isolation (i.e, in a managed quarantine facility, with zero transmission during this period) then a 10 day quarantine without a test to release averts 94% (95% UI: 87%, 98%) of transmission, which rises to 95% (95% UI: 89%, 99%) with an LFT test to release and 95% (95% UI: 91%, 100%) with a PCR test to release. The remaining transmission potential may occur during the flight, and through undetected infections, and can be eliminated with either a pre-flight LFT or PCR test (Figure 4). No additional transmission is averted through a longer quarantine period of 14 days.

Delays between the taking of pre-flight tests and the time of flight departure may introduce additional transmission risk in the destination country as individuals may become exposed in the interim, leading to less sensitive but rapid tests averting more transmission if they can be conducted immediately prior to the flight (Figure S4). For example, a 3 day delay from PCR test to flight (as required by many countries) may reduce *R*_*s*_ by 41% (95% UI: 14%, 63%) whereas an immediate pre-flight LFT may reduce *R*_*s*_ by 64% (95% UI: 43%, 89%).

### Number of infectious arrivals

The number of infectious arrivals to each country is a function of the prevalence in, and flight volumes from, all other countries and varies substantially across and within regions (Figure 5). Amongst the studied countries only Australia (AUS), China (CHN), Singapore (SGP), and Vietnam (VNM) are at very high risk (where, in the absence of any interventions aimed at restricting importation, infectious importations are more than 100% of domestic incidence), due to having extremely low incidence based on the available data.

**Figure 5:**
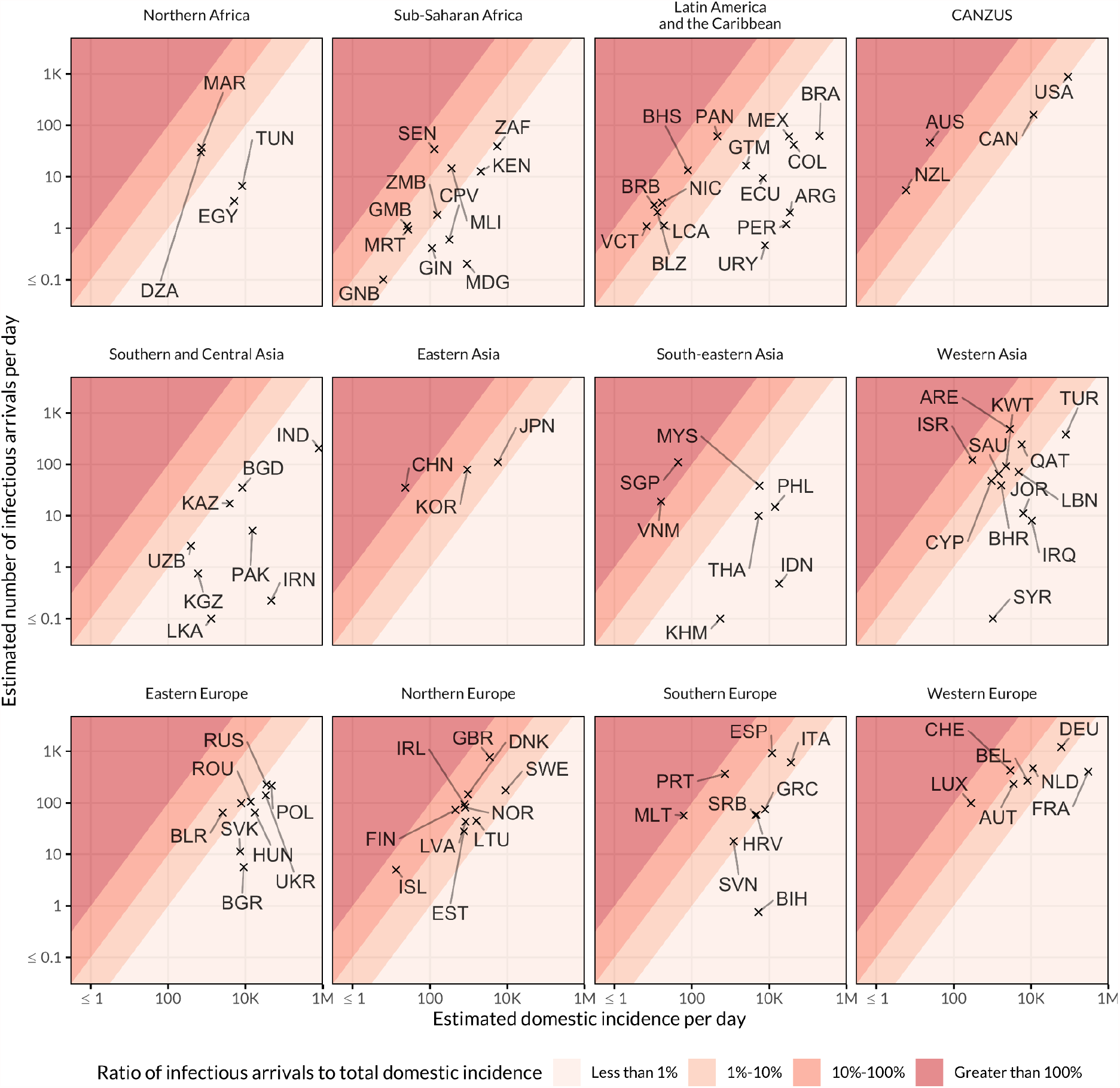
Estimated median risk of infectious imported cases. Risk is represented as the number of infectious imported cases per day as a percentage of domestic incidence in the destination country, as of 26 May 2021. Countries are grouped by UN region and subregion except for Northern America and Australia and New Zealand (CANZUS) and Southern Asia and Central Asia (Southern and Central Asia).

**Figure 6:**
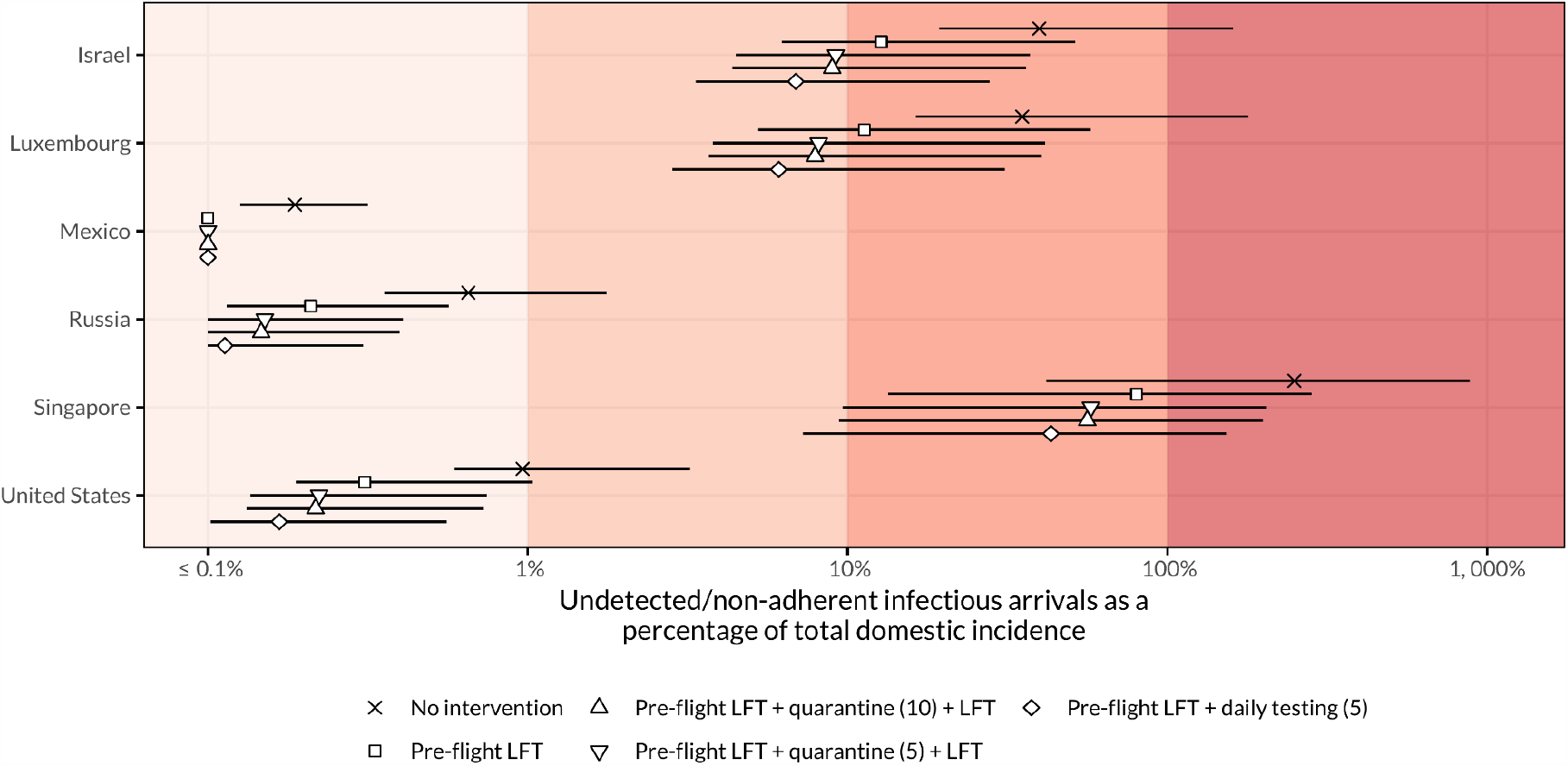
Effectiveness of four testing and/or quarantine strategies, compared to no intervention. Risk is derived as the ratio of new infectious arrivals to domestic incidence, expressed as a percentage. Results are shown for six selected countries for the following strategies in increasing order of reduction of entries: no intervention; pre-flight LFT with no further quarantine or testing; pre-flight LFT followed by five days of quarantine with an LFT at exit; pre-flight LFT with ten days of quarantine and an LFT at exit; pre-flight LFT followed by daily LFT for five days. Points represent median risk, with the horizontal line showing the 95% UI; where the median or endpoint of the UI is less than 0.1%, the value is shown as “≤0.1%”. Plots for all included countries may be found in the Supplementary appendix (Figure S5).

Given a current rate of importation of infections and domestic incidence, achieving a particular target risk level in a given country will require different effectiveness of interventions (Figure 4, Figure 6). For example, the introduction of daily tests for five days (with a pre-flight LFT) would reduce Luxembourg’s risk from a median of 35% (95%: 16%, 179%) to a median risk of 6% (95%: 3%, 31%). In contrast, the introduction of this strategy in Singapore would reduce its risk only as low as 43% (95%: 7%, 153%) as its baseline risk is among the highest in the world (249%, 95%: 42%, 882%) due to a large number of arrivals and low domestic incidence. Conversely, the United States’ high domestic incidence means the risk of importation with no intervention strategy is 1% (95%: 0.6%, 3.2%) indicating that restrictions on travellers may not be an effective way to prevent onwards transmission, even if the introduction of daily testing would reduce the median risk from importation to 0.2%.

### Variants of concern

Importation of the B.1.1.7 variant (first identified in the United Kingdom) has varied over time with incidence of the variant in the UK and travel from the UK to other countries (Figure 7). Importation as a percentage of domestic incidence is therefore dependent on the epidemic at both ends of the travel route and the proportion of UK cases which are the variant of concern, and hence many different importation risk profiles, over time, may be observed (solid circles in Figure 7). We consider the implementation of the intervention we specify as a strategy for comparison, a pre-flight LFT, five days of quarantine and LFT at completion (assuming adherence with isolation and quarantine guidance from published literature). Assuming LFT and PCR do not exhibit variant-specific sensitivity, such an intervention would reduce the risk of importation of infected travellers by 88% (95% UI: 72%, 94%) (open circles in Figure 7) and the risk of importation of infectious travellers by 84% (64%, 95%). Here we present results for infected arrivals, whether infectious or not, for Israel, Luxembourg, Mexico, Russia, Singapore and the United States of America.

**Figure 7:**
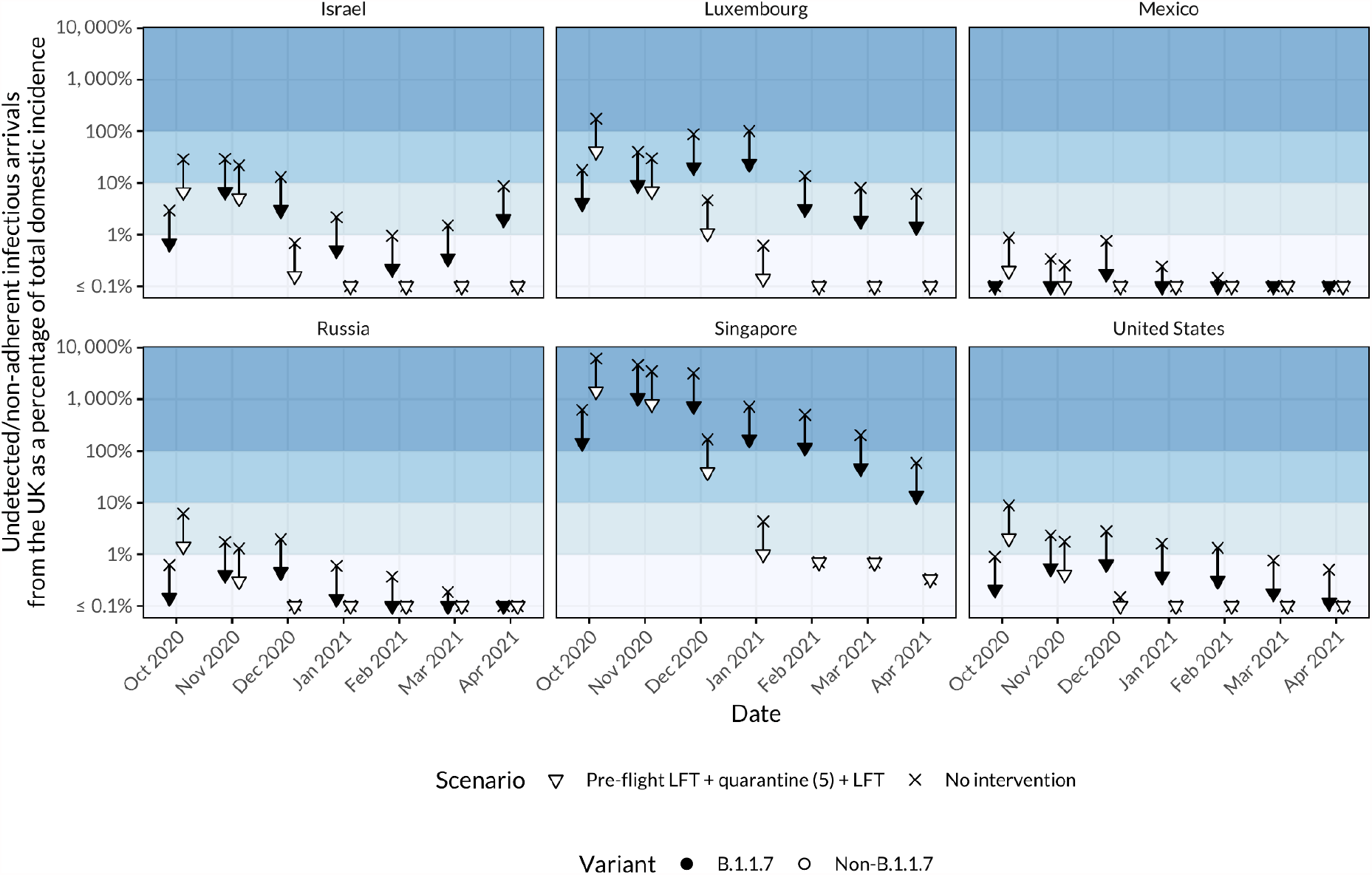
Median risk of importation of the B.1.1.7 (diamonds) variant of concern and non-B.1.1.7 (circles) variants from the United Kingdom as a proportion of total domestic incidence. Filled points indicate baseline rates of importation, and open points represent the impact of the intervention scenario (pre-flight LFT, 5 day quarantine with LFT at exit). The y axis is cropped at 0.1% although some very low risk months for some countries have both a baseline risk and reduced risk less than 0.1% (e.g. March 2021 in Mexico for both B.1.1.7 and non-B.1.1.7 with and without intervention).

At one extreme, Singapore has a small domestic epidemic (on the order of tens of cases per day) during the time October 2020 to March 2021. Importation of non-B.1.1.7 peaks in November 2020, approximately 62 times that of the total domestic incidence (95% UI: 57-fold, 68-fold) and B.1.1.7 importation peaks a month later in December, at approximately 46 times the total domestic incidence (95% UI: 42-fold, 51-fold). In contrast, the United States has incidence of the order 10,000s-100,000s and importation from the UK of B.1.1.7 peaks at a risk of only 3% (95% UI: 2%, 3%) of total domestic incidence in December 2020 with non-B.1.1.7 declining steadily from a peak of 9% (95% UI: 5%, 12%) of domestic incidence in October 2020 to less than 0.1% (95% UI: 0%, <0.1%) by December 2020. Similar patterns to the USA are seen in Mexico and Russia, where incidence was in the tens of thousands and peaked in either December 2020 (Russia) or January 2021 (Mexico). Israel’s risk of importation of B.1.1.7 from the UK has been increasing since January 2021 as its estimated domestic epidemic peaked at that time (8218, 95%: 7933, 9071) and the B.1.1.7 makes up a larger proportion of exported cases from the UK and there is an increasing amount of air travel from the UK to Israel over this time.

A table providing the total domestic incidence and risk of B.1.1.7 and non-B.1.1.7 importation for the countries in Figure 7 from October 2020 to March 2021 is provided in the Supplementary Appendix (Table S1).

## Discussion

Here we estimate the risk of SARS-CoV-2 case importation to 99 countries, and evaluate possible strategies to reduce onward transmission from those imported cases through quarantine and/or testing. Interventions enacted at the origin such as pre-flight testing may prevent the majority of infectious arrivals (pre-flight LFT and PCR preventing 66% (95% UI: 48%, 86%) and 85% (95% UI: 73%, 96%) respectively). We find that a requirement of strict isolation upon symptom onset, as is the current recommendation for any suspected case, will avert a substantial volume of post-flight transmission (reduction in *R*_0_: 45% (95% UI: 28%, 64%)). Furthermore, testing with LFTs or PCR after 5 days of quarantine (with release if negative) may match or exceed the effectiveness of the 14-day quarantine period in reducing transmission potential. Alternatively, daily LFTs upon arrival may avert a substantial amount of transmission while allowing for the avoidance of quarantine if negative. We find combined strategies involving both pre- and post-flight testing such as a pre-flight LFT combined with daily LFT tests for 5 days on arrival may prevent most infectious individuals from flying and avert transmission from those early in the incubation period who may go undetected by a pre-flight test (reduction in *R*: 91% (95% UI: 75%, 98%)).

We find that lateral flow tests may be a valuable tool to avert transmission from infected travellers, especially when used repeatedly over the course of 5 or 7 days. Their rapidity may also allow for their use immediately prior to boarding a flight, e.g. as at Stansted airport (19), whereas PCR tests would often require 24 hours to return results. There is evidence that transmission during a flight is possible (20) and that masks may be effective in reducing the risk of transmission (21). Immediate pre-flight rapid tests should be considered to reduce the in-flight transmission risk from currently infectious travellers in combination with the aforementioned measures. There has been concern over the sensitivity of LFTs when compared to the current gold standard PCR test, however they have been shown to detect culturable virus with 97% sensitivity (10), and those who are most likely to transmit to others with 83% sensitivity (22). In addition, while PCR tests are highly sensitive, their ability to detect residual non-viable RNA for several weeks following the infectious period may lead to the continued isolation and disrupted travel of individuals who are no longer infectious (23). Nonetheless, both PCR and LFT may fail to detect infections in the first days following exposure when viral loads are low (with PCR likely to detect infections earlier), leading to the lower estimated effectiveness of a single test either pre- or post-flight. However, despite high reported specificity of lateral flow tests (99.97% in the UK (24)) high volumes of testing will inevitably lead to false positives and, as such, the cost-effectiveness of confirmatory testing with a second lateral flow test or PCR, and potential non-independence thereof, should be assessed with a view to increasing the positive predictive value of testing (25). A further limitation of the reliance on culturability is that while a positive culture indicates sufficient whole virus to cause transmission, a negative culture may not necessarily imply an inability to infect. The relationship between Ct value, culturability and infectivity is key to characterising the ability of tests to detect infectious individuals and reliance on PCR is likely to detect and isolate some individuals well past the end of their infectious period.

Adherence to both quarantine or self-isolation after a positive test or symptom onset is a major component of the effectiveness of all post-flight intervention strategies explored and is one of the least-well characterised components of quarantine, testing and contact tracing systems. We use broadly indicative values for imperfect adherence to quarantine and self-isolation for contact-traced individuals due to a lack of data on rates of adherence of travellers in different countries over the course of the pandemic (26–29). In particular, there are likely cultural and social factors affecting adherence to quarantine and self-isolation (27) and we recognise that estimates of adherence from the UK and Norway may not be transferable to other countries. There is evidence that the rate of adherence to the 14-day quarantine in the absence of symptoms is low, at least in the UK where those self-isolating still report 3.85 (SD: 4.67) non-essential outside trips within the last week (26). However, adherence to self-isolation following a positive test is reportedly higher (86% (12)), although may be biased upwards as a result of this being a requirement under UK law. A recent ONS survey in the UK found that 17% of individuals reported non-adherence to self-isolation guidelines, namely leaving the home (83% of non-adherents) or having visitors for non-permitted reasons. 80% of individuals self-isolating in a household shared with others were unable to fully self-isolate (14) which poses a challenge where poor household isolation and potential low adherence to guidelines for household contacts may result in community transmission linked to an infectious traveller (30). Those who left the home chiefly did so to obtain essential supplies (32%) or to attend work or university (31%) and 30% of respondents who had tested positive either misunderstood or were unaware of the self-isolation requirements, indicating a need for targeted financial and social support, clearer guidance on how to self-isolate, e.g. staying in one room, wearing a mask in common areas (24), and strategies to reduce the risk of transmission under shorter quarantine periods that are more manageable for individuals (31).

There may be a trade-off in the effectiveness of quarantine between its duration and adherence in the population, as shorter quarantines may be easier to adhere to. Managed isolation of travellers in designated facilities such as hotels with regular visits from public health workers, as employed in East Asia and Oceania (32), may minimise possible transmission due to non-adherent persons. This strategy may be considered for countries in which the ratio of possible imported cases to domestic incidence is high (e.g. imported infections may make up the majority of new cases) and where there is a desire to exclude all variants of concern (in which case the threshold for action may be lower). Requiring managed quarantine in designated facilities may also prevent outbreaks within the household of the returning traveller, which are liable to spread further unless the entire household is required to quarantine.

Previous work by Russell and colleagues (3) suggests that an individual country’s risk from imported SARS-CoV-2 infections should be considered relative to their domestic incidence, with the required stringency of interventions being proportional to that risk. While this principle is still broadly applicable, the emergence of novel variants such as those recently detected in the UK (33,34), South Africa (35), Brazil (36) and India (37) with the potential for greater transmissibility, mortality, and potential for immune escape necessitate the stratification of importation risk and quantifying not just the number of arrivals who may spread the variant, but how many additional infections are likely to occur. In response, many countries have since implemented more stringent travel restrictions to prevent entry of variants. While we estimate that managed quarantine (with the assumption of complete self-isolation) may reduce the modelled risk to zero, real-world assessment of this policy found outbreaks may still occur (38), which indicates that even strict travel restrictions may be limited to the delay, but not prevention, of the importation of variants (39). If a given variant is already present, local interventions such as contact tracing (2) will be required to limit internal spread. For example, variants such as B.1.1.7 have been detected in many other countries outside of the UK and quickly became the dominant circulating strain in Europe and the USA (40). Testing of incoming travellers may be valuable as a surveillance tool to monitor the incidence of importation of variants; as LFTs detect only the nucleocapsid of SARS-CoV-2, positive lateral-flow tests should be followed up with PCR to monitor for S-gene target failure (for B.1.1.7) or to carry out further genomic analyses. Another factor is the current level of restrictions or population immunity (through infection or vaccination) in the destination country (3,41); imported cases arriving into an *R* < 1 environment will be much less likely to seed new local epidemics than in a *R* > 1 environment.

A limitation of this work, in common with other studies relying on infection fatality ratios to estimate the level of under-ascertainment of cases or infections globally (42), is that under-ascertainment and under-reporting of COVID-19 deaths is also known to be occurring at significant levels in numerous countries (43). Many countries’ estimates of the prevalence and incidence of COVID-19 are known, or highly suspected, to be biased downwards, due to death under-ascertainment. This means that any country that is suspected to be underestimating mortality rates of COVID-19, is likely to also underestimate the prevalence and incidence of infection, if such estimates are arrived at using modelling frameworks fit to mortality data — the method used here and the method most global modelling efforts use. Improving global estimates using serological and other surveillance data is ongoing but extremely challenging, given data availability issues and the differences in data quality between different countries. Tools such as Serotracker (44), OpenSky (15) and existing excess deaths databases (45) could provide the tools to arrive at global estimates of death under-ascertainment; but are outside the current scope of this study. The routine testing of travellers, as explored in this study, would allow for the estimation of prevalence in other countries after adjustment for factors such as travel volume.

For clarity and brevity, we have presented the risk to a given country for all incoming travel, and not from specific individual countries. Some countries have chosen to impose restrictions on flights from high-risk origin countries, and relax restrictions on flights from countries considered low-risk in “travel corridors” or “air bridges” (46). Such a strategy is problematic due to the potential for multi-leg flights and mixing with others from high-risk countries in tourist areas of an intermediate country. To allow for country-specific estimates to be calculated, we present relative measures of reduction in infectious entries and their transmission potential by each strategy so that if prevalence and travel volumes from a specific country are known, absolute risk may be simply calculated as the product of prevalence, travel volume and relative reduction. It should be noted that our absolute risk estimates are based on prevalence and travel volume as of April 2021, and that assessing risk in terms of infectious entries may underestimate the effectiveness of quarantine and testing programmes.

In this report we have shown that existing strategies to reduce SARS-CoV-2 importation such as a 14 day quarantine period for arrivals are effective at reducing risk, and that the duration of quarantine may be reduced to 10 days without, and 5 days with, a PCR or rapid lateral flow antigen test to exit quarantine if negative. Additionally, 5 days of lateral flow tests taken daily could allow for the removal of mandatory quarantine, even under less than perfect adherence. Requiring pre-flight tests as close to departure as possible (i.e, an advantage of rapid tests) may prevent the majority of transmission from infectious would-be travellers. Our findings align with the findings of several other modelling studies for reducing the duration of quarantine in air travel and contact tracing such as Wells et al. (47) and Ashcroft et al. (48). All strategies are however highly dependent on the rate of adherence to quarantine and self-isolation, and improving these rates through financial and social support, and clarity of guidance, will be key to the success of such strategies (27). Managed quarantine on arrival can help minimise the risk of importation of variants of concern from high risk destinations. The risk of infectious arrivals causing ongoing transmission in a given country should be considered relative to domestic incidence (and domestic R), with restrictions on travel having a higher relative impact in countries where the expected number of infectious arrivals exceeds domestic incidence, where a large proportion of the population remains susceptible, and where there is a desire to exclude the importation of variants of concern. Travel restrictions carry significant economic, political, and social costs which must be weighed against the contribution of imported cases to SARS-CoV-2 incidence.

## Data Availability

Code is available at https://github.com/cmmid/covid_quar_test_import_risk for the analysis conducted within this article.

## Data Availability

After publication, the data will be made available upon reasonable requests to the corresponding author. A proposal with detailed description of study objectives and the statistical analysis plan will be needed for evaluation of the reasonability of requests. Deidentified data will be provided after approval from the corresponding author and the Mayo Clinic.

## Acknowledgements

The following funding sources are acknowledged as providing funding for the named authors. This research was partly funded by the Bill & Melinda Gates Foundation (OPP1139859: BJQ). This project has received funding from the European Union’s Horizon 2020 research and innovation programme - project EpiPose (101003688: WJE). This research was partly funded by the National Institute for Health Research (NIHR) using UK aid from the UK Government to support global health research. The views expressed in this publication are those of the author(s) and not necessarily those of the NIHR or the UK Department of Health and Social Care (16/136/46: BJQ; 16/137/109: BJQ; PR-OD-1017-20002: WJE). UK MRC (MC_PC_19065 - Covid 19: Understanding the dynamics and drivers of the COVID-19 epidemic using real-time outbreak analytics: SC, WJE). Wellcome Trust (206250/Z/17/Z: TWR; 208812/Z/17/Z: SC, SFlasche). This publication has been supported by German Federal Ministry of Health (BMG) COVID-19 Research and development funding to WHO.

The following funding sources are acknowledged as providing funding for the working group authors. This research was partly funded by the Bill & Melinda Gates Foundation (INV-001754: MQ; INV-003174: KP, MJ, YL; INV-016832: SRP; NTD Modelling Consortium OPP1184344: CABP, GFM; OPP1191821: KO’R). BMGF (INV-016832; OPP1157270: KA). CADDE MR/S0195/1 & FAPESP 18/14389-0 (PM). EDCTP2 (RIA2020EF-2983-CSIGN: HPG). ERC Starting Grant (#757699: MQ). ERC (SG 757688: CJVA, KEA). This project has received funding from the European Union’s Horizon 2020 research and innovation programme - project EpiPose (101003688: AG, KLM, KP, MJ, RCB, YL). FCDO/Wellcome Trust (Epidemic Preparedness Coronavirus research programme 221303/Z/20/Z: CABP). This research was partly funded by the Global Challenges Research Fund (GCRF) project ‘RECAP’ managed through RCUK and ESRC (ES/P010873/1: CIJ). HDR UK (MR/S003975/1: RME). HPRU (This research was partly funded by the National Institute for Health Research (NIHR) using UK aid from the UK Government to support global health research. The views expressed in this publication are those of the author(s) and not necessarily those of the NIHR or the UK Department of Health and Social Care200908: NIB). MRC (MR/N013638/1: EF; MR/V027956/1: WW). Nakajima Foundation (AE). NIHR (16/137/109: FYS, MJ, YL; 1R01AI141534-01A1: DH; NIHR200908: AJK, LACC, RME; NIHR200929: CVM, FGS, MJ, NGD; PR-OD-1017-20002: AR). Royal Society (Dorothy Hodgkin Fellowship: RL). Singapore Ministry of Health (RP). UK DHSC/UK Aid/NIHR (PR-OD-1017-20001: HPG). UK MRC (MC_PC_19065 - Covid 19: Understanding the dynamics and drivers of the COVID-19 epidemic using real-time outbreak analytics: NGD, RME, YL; MR/P014658/1: GMK). UKRI (MR/V028456/1: YJ). Wellcome Trust (206250/Z/17/Z: AJK; 206471/Z/17/Z: OJB; 210758/Z/18/Z: JDM, JH, KS, SA, SFunk, SRM; 221303/Z/20/Z: MK). No funding (DCT, SH).

## Supplementary appendix

**Figure S1:**
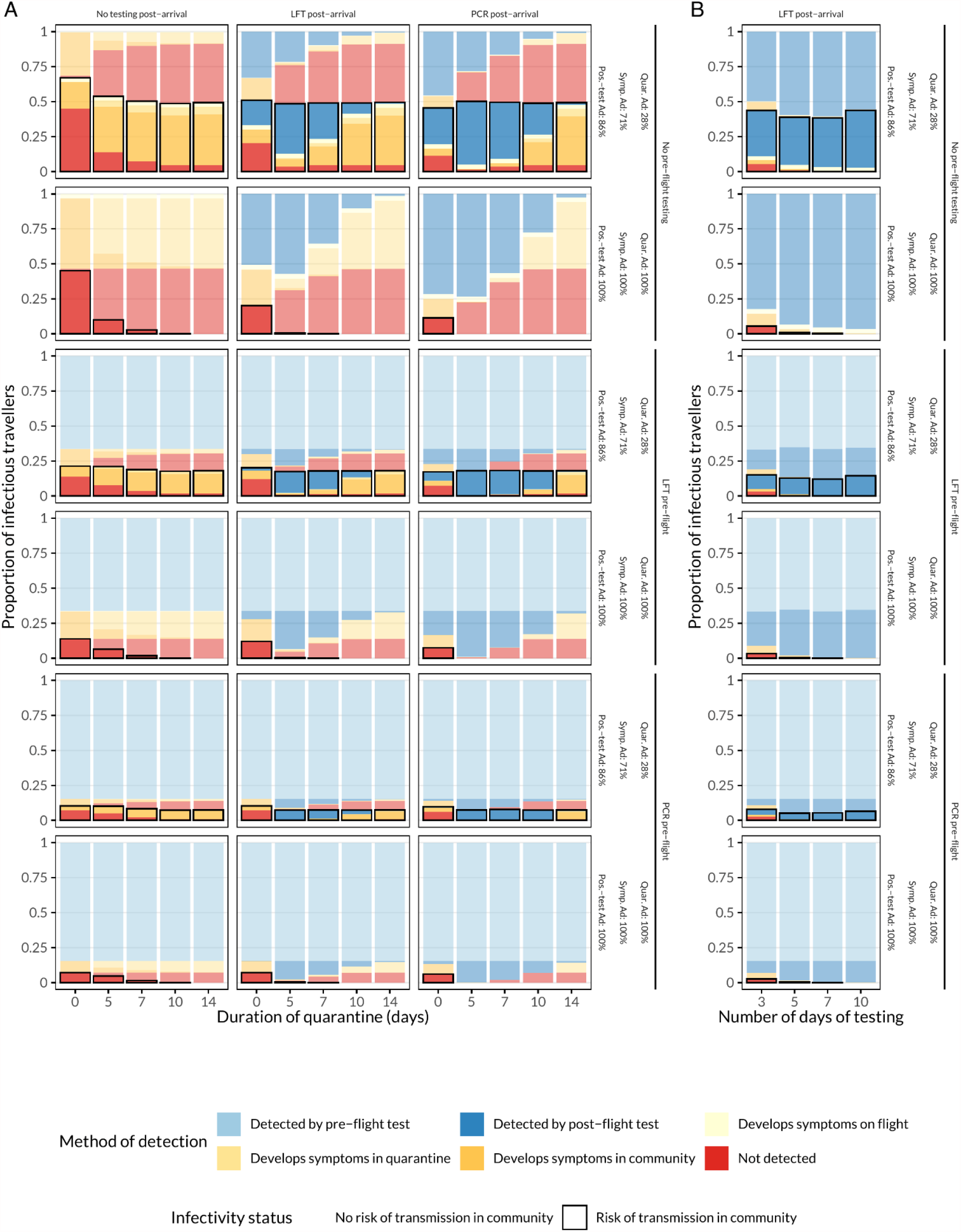
Mean proportion of intending travellers detected at each stage of the quarantine and testing strategies (columns corresponding to either A) quarantine with either no test, Lateral Flow test (LFT), or Polymerase Chain Reaction (PCR) test or B) daily testing). Rows in each plot correspond to either no pre-flight testing, or pre-flight testing with LFT or PCR and either perfect adherence to quarantine and self-isolation guidance, or values derived from literature.

**Figure S2:**
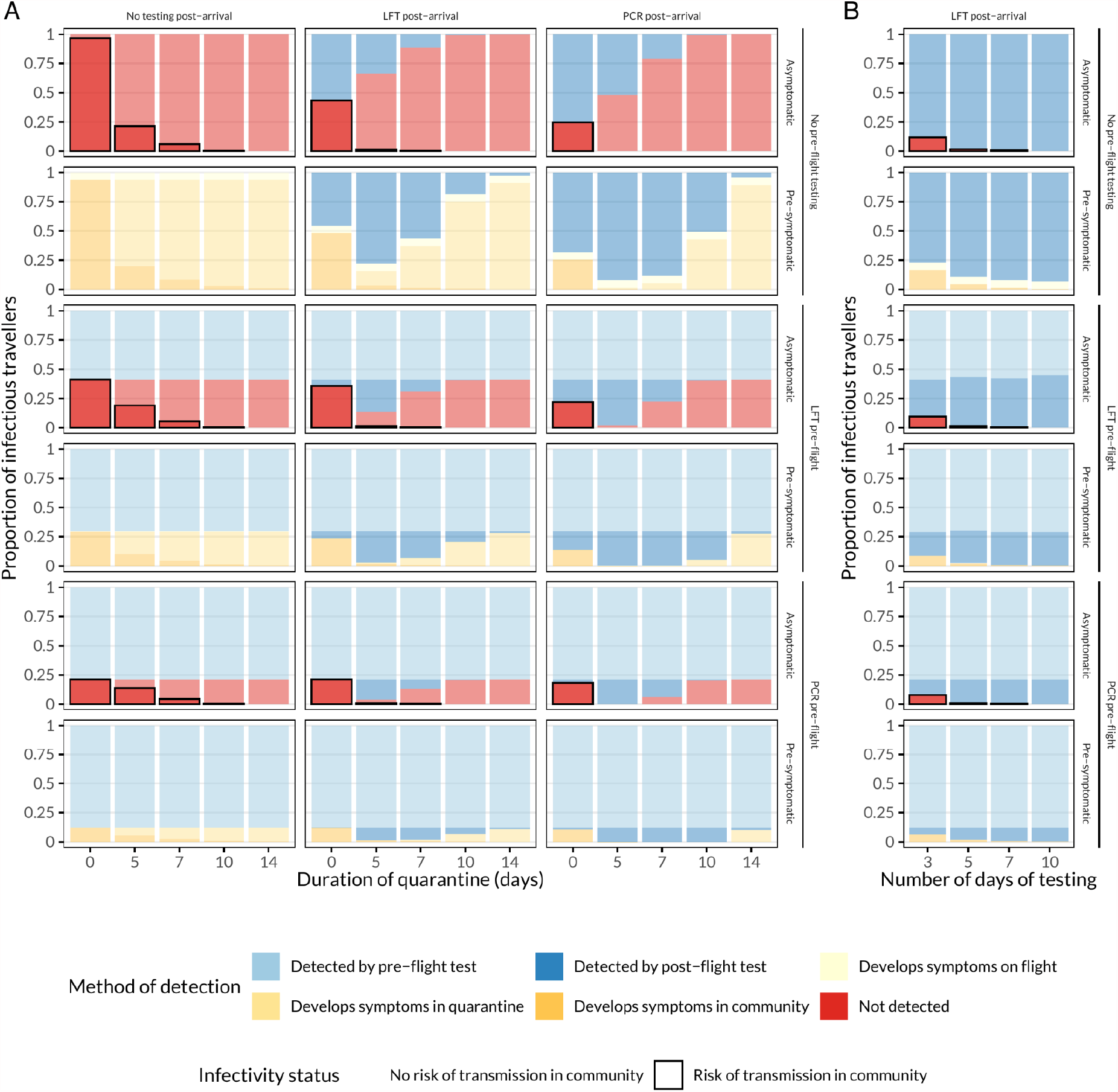
Mean proportion of intending infectious travellers detected at each stage of the quarantine and testing strategies, stratified by whether or not individual is ever symptomatic or always asymptomatic (columns corresponding to either A) quarantine with either no test, Lateral Flow test (LFT), or Polymerase Chain Reaction (PCR) test or B) daily testing). Rows in each plot correspond to either no pre-flight testing, or pre-flight testing with LFT or PCR and either perfect adherence to quarantine and self-isolation guidance, or values derived from literature.

**Figure S3:**
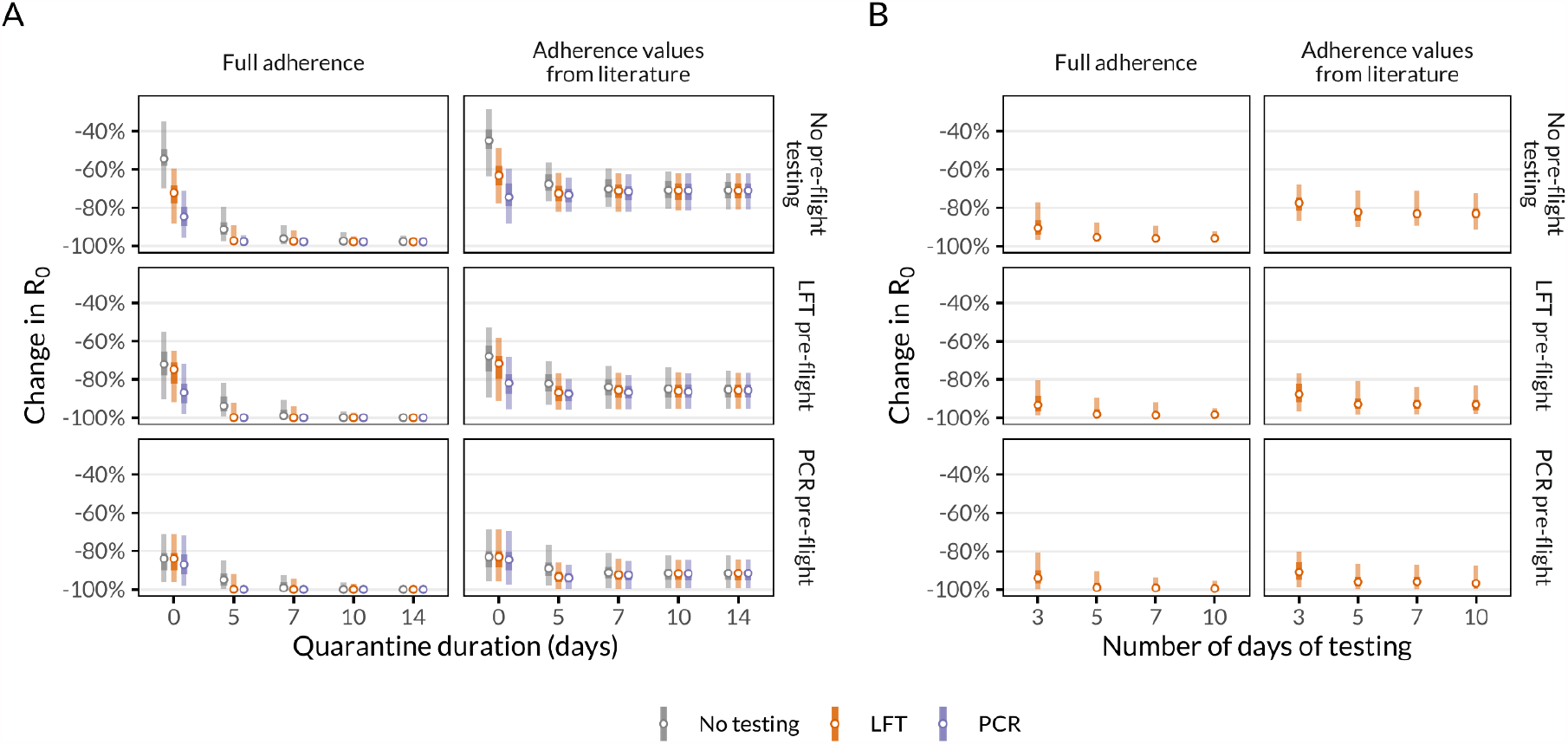
Change in *R*_*0*_ of infectious arrivals entering the community, including the effect of symptomatic self-isolation. Self-isolation only with full adherence (top row of plots) or adherence values from literature (28% of individuals adhering to quarantine, 71% of individuals adhering to post-symptom onset self-isolation, and 86% adhering to post-positive test isolation, bottom row of plots), and with or without pre-flight tests. A) Quarantine of varying durations with or without testing with LFTs and PCR. B) Daily testing without quarantine with lateral-flow tests, with self-isolation only upon a positive test result. Vertical lines represent 95% (outer) and 50% (inner) uncertainty intervals around medians (points). Note discrete x-axis values for quarantine duration and number of days of testing.

**Figure S4:**
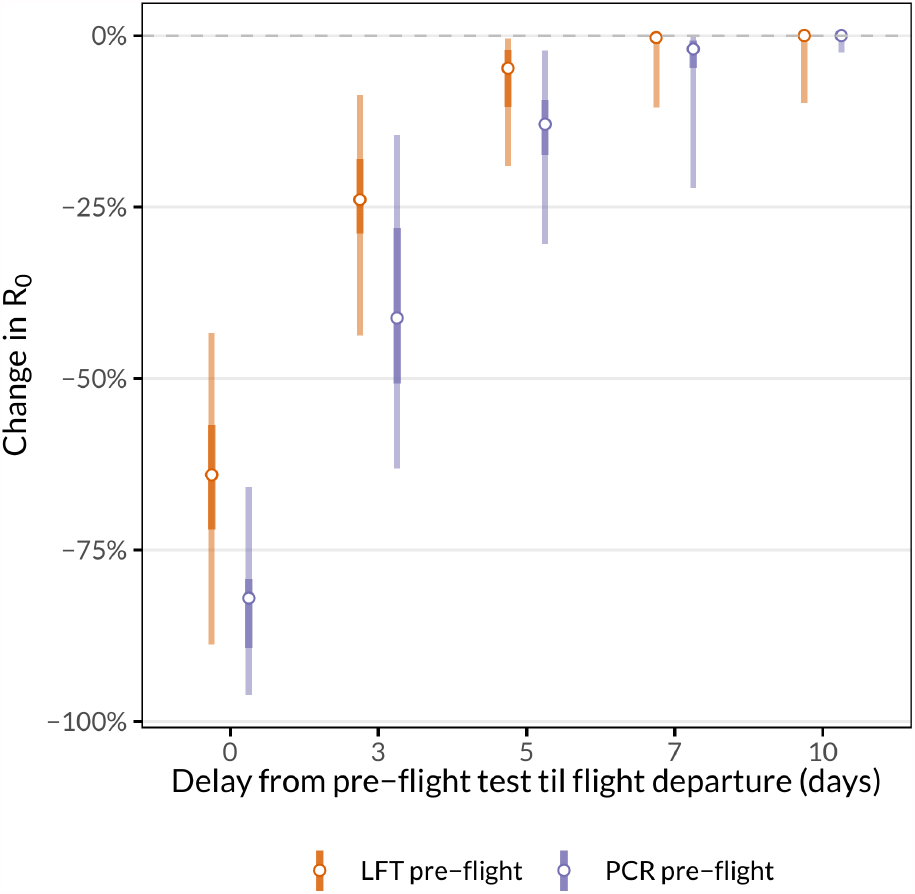
Change in *R*_0_ of infectious arrivals with different delays from a pre-flight test until boarding a flight. Vertical lines represent 95% (outer) and 50% (inner) uncertainty intervals around medians (points).

**Figure S5:**
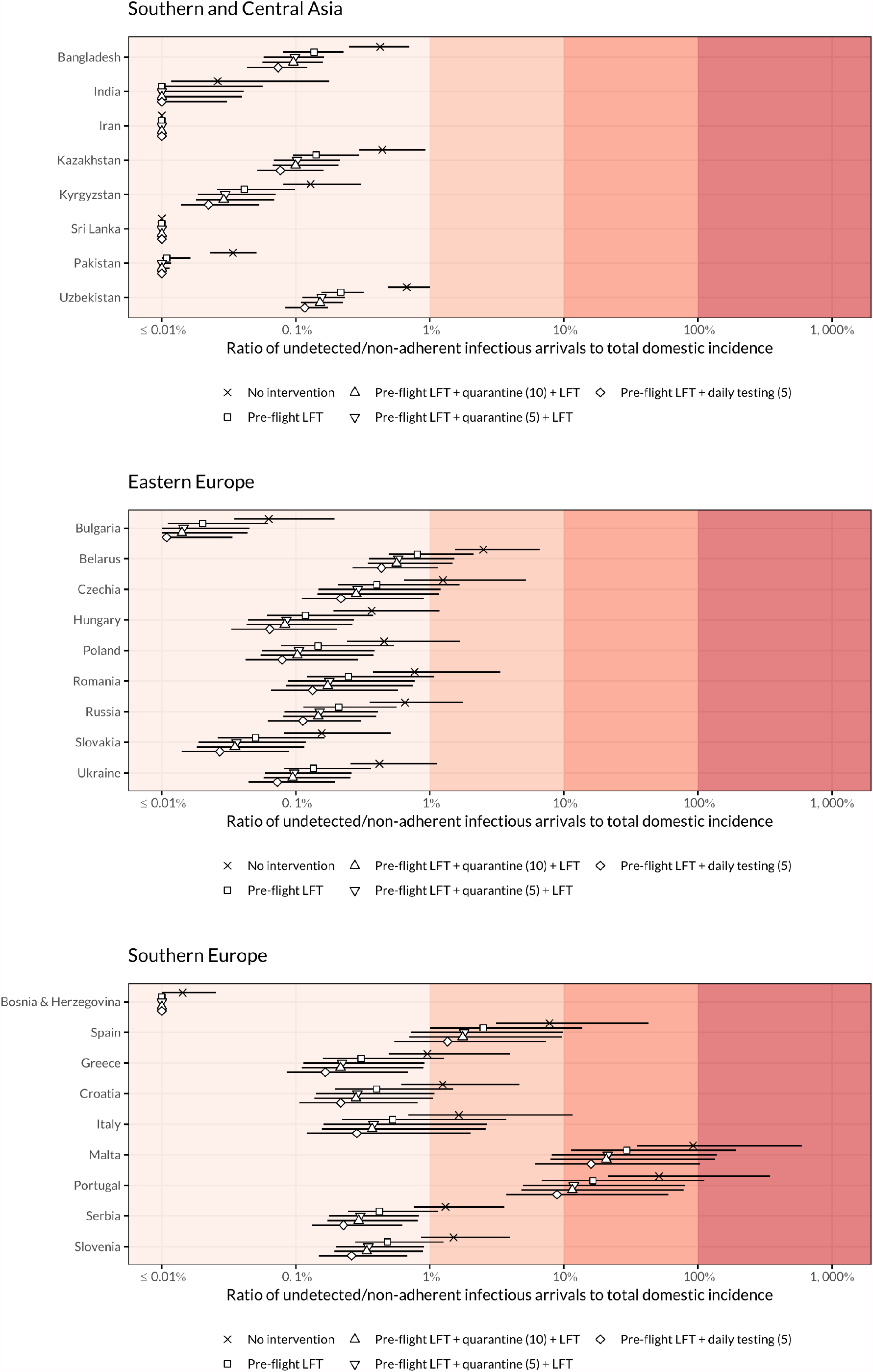
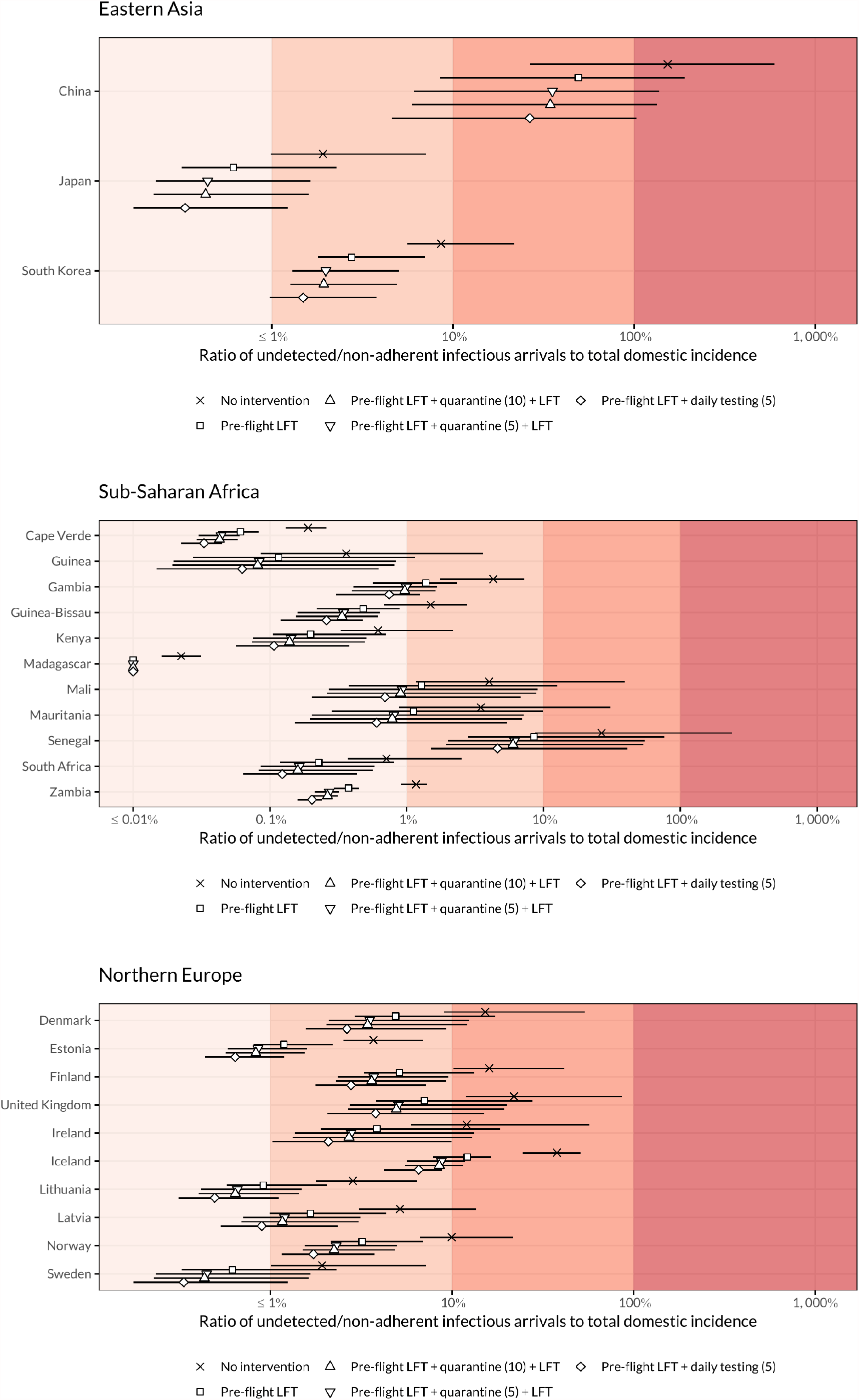
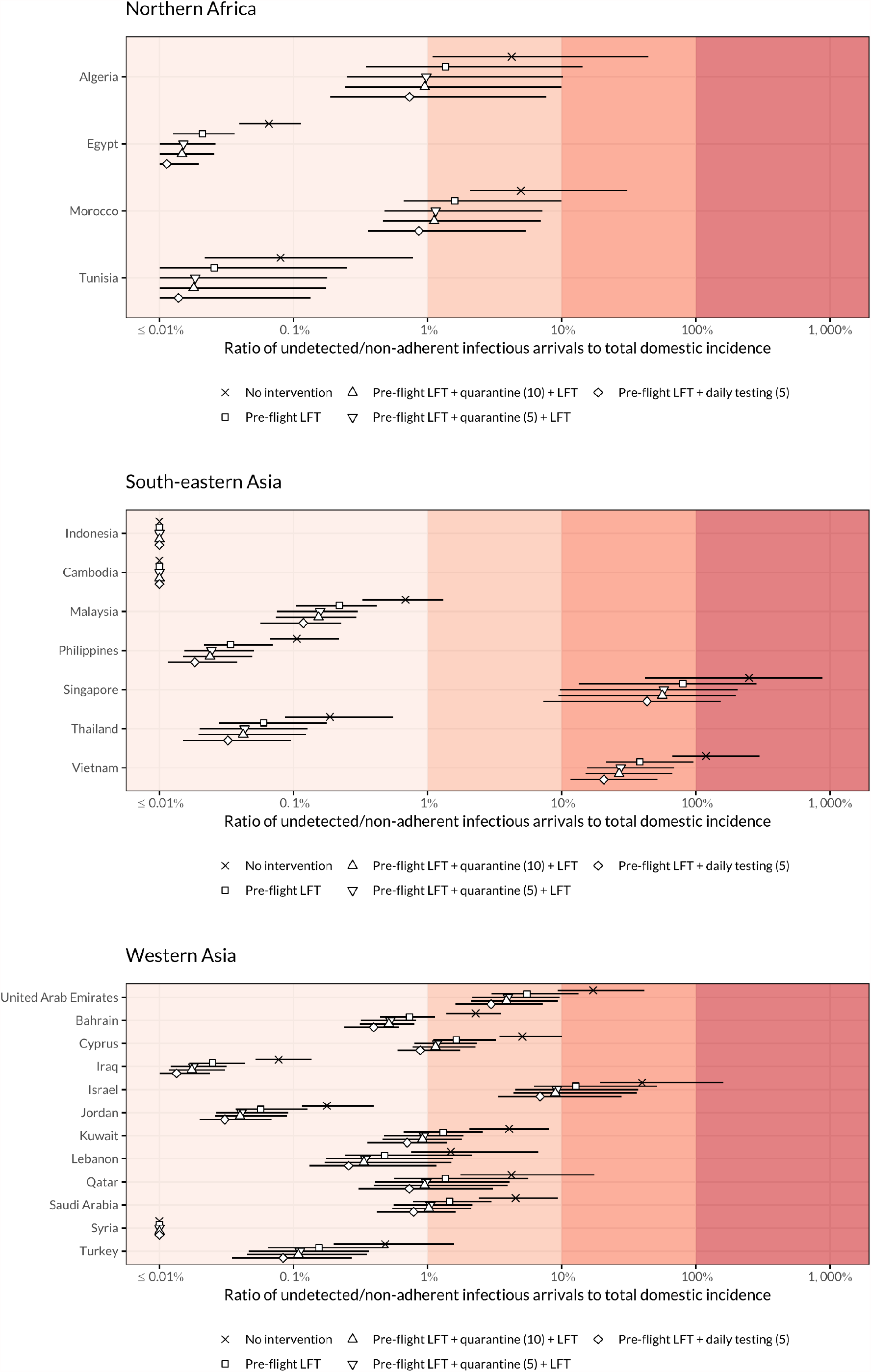
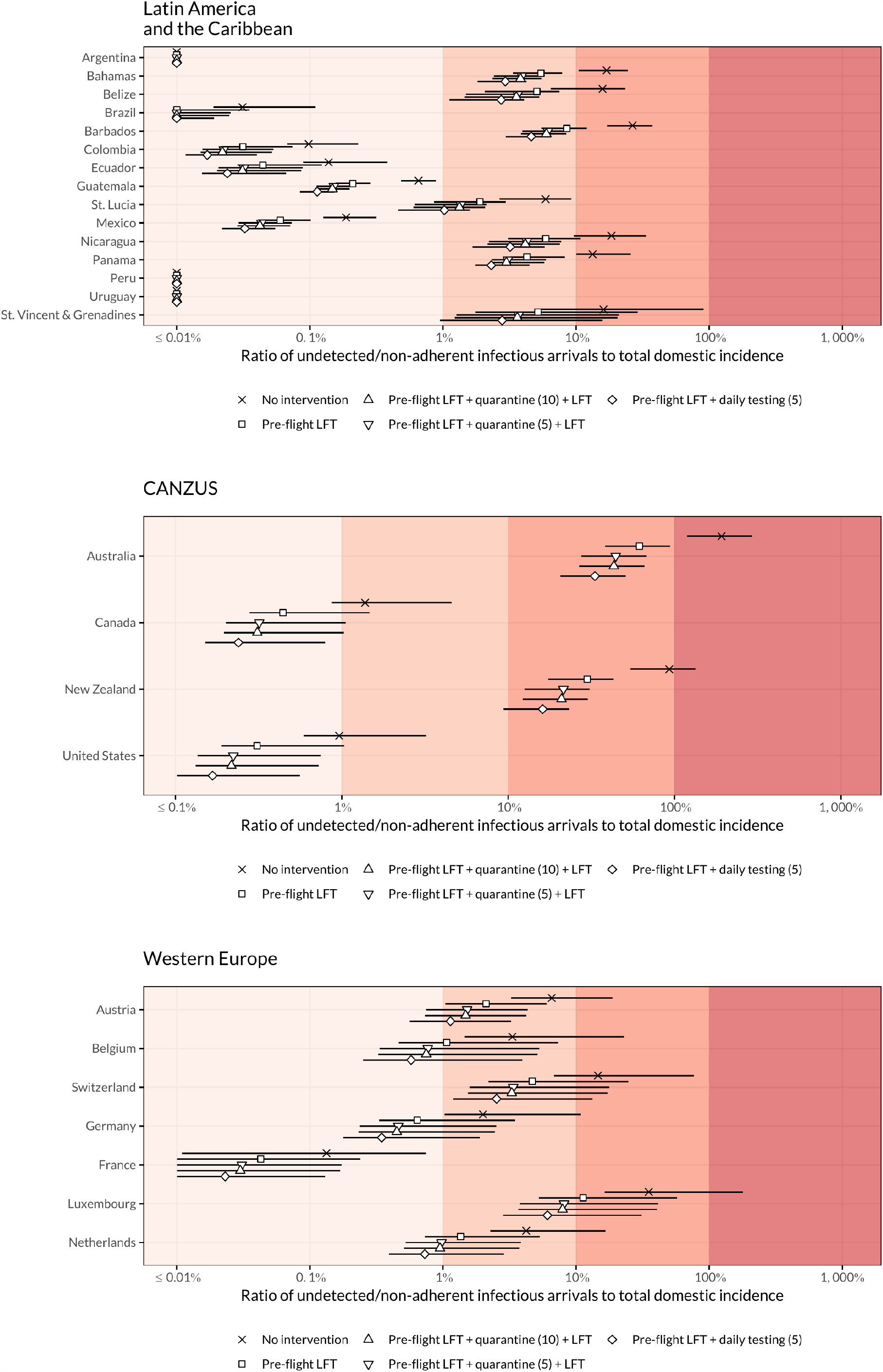
Effectiveness of four testing and/or quarantine strategies, compared to no intervention. Risk is derived as the ratio of new infectious arrivals to domestic incidence, expressed as a percentage. Results are shown for all included countries for the following strategies in increasing order of reduction of entries: no intervention; pre-flight LFT with no further quarantine or testing; pre-flight LFT followed by five days of quarantine with an LFT at exit; pre-flight LFT with ten days of quarantine and an LFT at exit; pre-flight LFT followed by daily LFT for five days. Points represent median risk, with the horizontal line showing the 95% UI; where the median or endpoint of the UI is less than 0.1%, the value is shown as “≤0.1%”.

### Detailed methodology: estimating time-varying under-ascertainment rates each day, for each country

We estimate prevalence and incidence for each country (with greater than 10 deaths in total). To do so, we estimate the level of under-ascertainment of symptomatic cases according to the methods in (42) within a fully Bayesian framework. The result of the inference is a time-dependent posterior distribution, representing the level of case ascertainment for each country. We then adjust the confirmed cases for each country using the median of the posterior distribution on each day, and the lower and upper 95% credible intervals. This process results in a 95% credible interval of the true number of symptomatic cases for each country. When considering all infections and not just symptomatic cases, we perform a final step adjusting for potential asymptomatic and presymptomatic infections. We assume that a large range (between 26% and 27%) of infections are asymptomatic (8).

To estimate the proportion of symptomatic cases ascertained over time, we fit a Gaussian process to a statistical Bayesian model for daily new deaths. The likelihood of the model, written in its simplest form, is given by

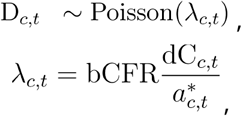

where D_*c, t*_ is the number of daily deaths for country on day. We assume a Poisson observation process, with a rate given by λ_*c, t*_, the product of the assumed true baseline case fatality ratio and the total number of cases with a known outcome by day *t*. The true number of cases is given by “adjusting” the ascertained number of cases dC_*c, t*_ with the ascertainment rate 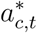. Specifically, the ratio of the two gives the true number of symptomatic cases in country *c* on day *t*. With the ascertainment rate defined in the likelihood function as a parameter, we are able to use the confirmed death data to fit our model and infer a time-dependent posterior distribution for this parameter.

The time-dependent ascertainment rate is defined as

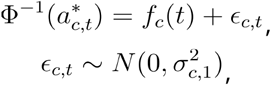

where *f*_*c*_(*t*) is a nonparametric function of time for country *c*, ∈ _*c, t*_ are independent normally distributed random variables to attempt to explain daily variation in ascertainment for country and finally Φ^-1^(*x*) is the inverse of the probit function mapping the ascertainment rate to the unit interval - the range of supported values of the ascertainment rate. We model *f*_*c*_(*t*) as a realisation of a univariate zero-mean Gaussian process:

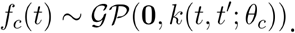

The details of this Gaussian process, for example the specific parameterisation of the covariance matrix and the kernel function and the priors used can be found in the study which originally developed this model (42).

### Adjusting for under-ascertainment

Firstly, we impute corresponding dates to the ascertainment estimates for each country. We do so by assuming the delay from confirmation to death follows the mean of an estimated distribution from the literature of 13 days (49). We have, at this stage, effectively produced a time series of daily ascertainment rates, if we consider only the median and the lower/upper 95% credible intervals of the posterior distribution. Finally, we adjust the confirmed cases on each day using the ascertainment estimates.

### Estimating infections

We estimate the total number of infections from the adjusted symptomatic case curves for each country (adjusted for under-ascertainment) by inflating them using estimates from a systematic review of the number of asymptomatic infections overall. The range given is 26% - 37% (8).

### Incidence and prevalence estimates

To estimate incidence for each country, we calculated the mean number of infections over the same time period as the time period considered for the expected number of imported cases (which depends on the specific scenario). This time period is typically either a week or a month depending on what exactly is being considered. However, our inference framework provides us with a crude incidence estimate for each country on each day. Therefore, we are able to perform ad-hoc calculations within the same framework over arbitrary time periods, if the traveller data used to estimate expected numbers of imported cases is over a different time period or of a different temporal resolution. To estimate prevalence, we use cumulative incidence, summed over the mean (10 days) of a distribution of the infectious period (50), as a proxy for prevalence.

### Sources of uncertainty

Several sources of uncertainty are captured in our final uncertainty range:

- the inferred infectious period, with an uncertainty range reported in Table 1 of the main text of Russell et al. (2020) (3).
- the assumed proportion of asymptomatic infections, with an assumed range of [26%, 37%].
- the confirmation-to-death distribution, with an uncertainty range with a 95% CI of (8.7, 20.9) that we integrate over in the Gaussian process fitting procedure (49).

### Limitations of our methods

We summarise the limitations of the original study here briefly and we discuss the limitations of the additional steps - extending the methods in (42) - employed in this study to arrive at prevalence estimates in detail. We do so, as the original study (42) which develops and describes the under-ascertainment model includes a verbose description of the limitations of the methods, up to the point of estimating incidence, in the Discussion section of the main text. Furthermore, the original study goes into more detail about such limitations in its Supplementary Material.

#### Estimating under-ascertainment

In order to estimate under-ascertainment in a flexible manner, we assume a global baseline severity of COVID-19 of 1.4%, with the range 1.1% – 1.7% comprising the standard deviation of a normally-distributed prior on the baseline CFR. It is known that CFR of COVID-19 varies between locations. However, given that our analysis is on the scale of countries, and the uncertainty in the estimate is included in the final 95% credible intervals of our reported results (along with other sources of uncertainty), the effects of the assumption are relatively minor. We do however perform an additional sensitivity analysis in the original study (42), whereby we adjust the baseline CFR value for each country based on the underlying age-distribution of each country, using age-stratified CFR estimates (51). In doing so, we test the sensitivity of the model to the assumed CFR value. We find that our conclusions are broadly unchanged, and our cumulative incidence estimates are in good agreement with available seroprevalence results (42). For other limitations of these estimates, please refer to the main text and supplementary material of the original study (42).

#### Estimating incidence and prevalence

Extending the methods of (42) — whereby the resulting outputs of the mathematical model are posterior distributions for adjusted incidence over time for all countries (adjusted for under-ascertainment) — to arrive at prevalence estimates adds some limitations to the final estimates. The most pertinent of which is the additional assumptions about timing. Given that the outputs of the original model take the form of incidence measurements, and our estimates are on the scale of countries, whereby estimates are bound to be crude for a multitude of reasons, we use cumulative incidence as a proxy measure for prevalence. To do so, we sum the recent incidence levels over the mean of an estimated distribution for the time-to-infectiousness and infectious periods (which sum to 10 days (50)) to arrive at prevalence estimates. We include the time-to-infectiousness distribution to allow for some level of presymptomatic transmission (50).

Incorporating these distributions into the otherwise fully Bayesian framework would alleviate this as a limitation of our study. However, in doing so, some of the desirable scalability and flexibility of the model as it stands would be lost, as additional assumptions about recovery and death rates would be required, which have been shown to vary significantly globally. In an attempt to keep the analysis scalable and parsimonious, applied in the same way globally, we opt for the simple adjustment to arrive at prevalence. In doing so, we are producing relatively crude estimates. However, we believe that the uncertainty included in the model as to the true proportion of asymptomatic infections – the source of most of the uncertainty in the 95% lower and upper credible intervals of the results reported – overshadows any additional minor error introduced by using cumulative incidence over the infectious period as a proxy for prevalence.

